# Detection of airborne respiratory viruses in pediatric patient rooms in Guangzhou, China

**DOI:** 10.1101/2023.02.23.23286335

**Authors:** Yanmin Xie, Eunice Yuen Chi Shiu, Dan Ye, Wenjie Zhang, Wenbo Huang, Zifeng Yang, Benjamin John Cowling, Nancy Hiu Lan Leung

**Author notes:** ZY and BJC should be considered joint corresponding authors. **Corresponding author:** Zifeng Yang, State Key Laboratory of Respiratory Disease, National Clinical Research Center for Respiratory Disease, First Affiliated Hospital of Guangzhou Medical University, 151 Yanjiangxi Road, Yuexiu District, Guangzhou, Guangdong 510120, China. **Alternate corresponding author:** Benjamin J. Cowling, School of Public Health, Li Ka Shing Faculty of Medicine, The University of Hong Kong, 7 Sassoon Road, Pokfulam, Hong Kong. These authors contributed equally to this work (Yanmin Xie, Eunice Yuen Chi Shiu, Dan Ye).

## Abstract

**Background:** Despite the controversy of aerosol transmission for different respiratory viruses, there are few direct comparisons. Respiratory virus detection in the air can inform transmission risk assessment in healthcare settings. We aimed to identify five common respiratory viruses in the air in pediatric patient rooms.

**Methods:** We sampled air with two-stage cyclone samplers continuously for 4 hours in 5-bed pediatric patient rooms in a tertiary hospital in China. Respiratory virus RNA/DNA recovered in the air were quantified by PCR.

**Results:** We conducted air sampling on 44 occasions from December 2017 through January 2020, and identified 24, 18, 16, 4 and 8 occasions which had ≥1 patient in the room tested positive for respiratory syncytial virus (RSV), adenovirus (AdV), parainfluenza virus (PIV), influenza B or A virus, respectively. Detection of influenza A viral gene copies was most frequent even when there were no known infected patients in the room (72%). Influenza B, AdV and RSV were detected in low to moderate frequencies, whether there were infected patients in close proximity (13-50%) or not (12-25%). PIV was rarely detected even when air samplers were placed in close proximity to infected patients (8%). About 10^3^–10^5^ copies/m^3^ were detected for all detected respiratory viruses.

**Conclusions:** Healthcare workers and visitors likely have substantial exposure to various respiratory viruses including influenza A/B viruses, RSV and AdV in pediatric patient rooms, even in the absence of infected individuals in close proximity, suggesting the potential value of improving indoor ventilation or air disinfection in hospitals.

## INTRODUCTION

Acute respiratory infections are a leading cause of morbidity and mortality in children globally, with the burden disproportionately falling in lower-income locations [1]. Common respiratory virus infections in children include respiratory syncytial virus (RSV), adenovirus (AdV), parainfluenza virus (PIV), influenza A virus and influenza B virus [2,3]. Respiratory virus infections have been thought to transmit via respiratory droplets of various sizes including fine particle aerosols. Contact and droplet precautions are currently recommended for most common respiratory viruses in healthcare settings [4]. The potential need for aerosol precautions, however, remains controversial for influenza virus [5,6] and has lacked attention for other respiratory viruses [7,8]. Epidemiologic studies have reported the molecular detection of RSV in the air both in droplets [9,10] and fine particles [11,12]. Few studies have investigated the possibility of airborne AdV [13-16] and PIV [17,18] transmission in healthcare settings. Furthermore, similar studies were usually conducted in single-bed or two-bed wards in higher income locations [12,13,19], and few were conducted in hospitals [20]. Given heighted awareness of the potential of other respiratory viruses transmitting through the airborne route [4,21], here we report an extension of our previous study on influenza [22] to other respiratory viruses, to quantify the detection and concentration of virus-laden particles and factors of contribution in the air. This study was conducted prior to the COVID-19 pandemic.

## METHODS

### Study design

The study was conducted in a tertiary hospital in Guangzhou, China from December 2017 through January 2020, using different inclusion criteria but similar air sampling and sample processing procedures as described in our previous study [22]. All pediatric patients (≤14 years old) were screened routinely by reverse transcription polymerase chain reaction (PCR) for respiratory virus infection using throat swabs at admission. An air sampling occasion was triggered when a patient room had at least one patient (“target” patient) who was admitted for acute respiratory symptoms and was PCR-positive for influenza B virus, RSV, AdV or PIV infection. Influenza A virus was not considered when triggering air sampling to increase the chance of identifying other respiratory virus infections, although the samples we collected were still tested for influenza A virus. If more than one patient was identified on the same day, patients with a higher body temperature measured at admission or with a more recent symptom onset was selected.

We then conducted continuous air sampling for 4 hours, by setting two two-stage cyclone air samples developed by the US National Institute of Occupational Safety and Health (NIOSH) in the room [23]. One sampler was placed as close to the bed of the target patient as possible near the patients’ head (‘closer’ NIOSH sampler), and the other was placed close to the patient next to the target patient further away from the bed (‘further’ NIOSH sampler). The NIOSH samplers were fixed 1.3m from the floor so that they were the same height as the children’s head when sitting on the bed. The NIOSH samplers collected air particles in 3 different size-fractions of 1µm, 1-4µm and >4µm at a flow rate of 3.5 L/min. Other information including basic environmental parameters (temperature, relative humidity and CO_2_), the number of visitors and health care workers visiting the room, door/windows opening situation, and nebulizer therapy conducted while sampling was collected at the start and again after 2 hours and 4 hours.

### Ethical approval

The Institutional Review Board of the University of Hong Kong, and the First Affiliated Hospital of Guangzhou Medical University approved this study. All patients and parents or legal guardians provided verbal informed consent. Written consent was deemed unnecessary, given that the study involved environmental air sampling and information on patients’ diagnoses was collected anonymously with no personal identifying information.

### Laboratory methods

RNA or DNA from the respiratory virus in air samplers was extracted by QIAamp RNA Mini Kit or QIAamp Viral RNA Mini kit (Qiagen Co. Ltd., Shanghai, China) using the manufacturer’s protocols [24]. Five respiratory viruses, including influenza A virus, influenza B virus, RSV, AdV and PIV, were tested by real-time PCR using Taqman designed primers and probes, TaKaRa reagent and protocol as previously described [25]. We considered samples with a clear reaction signal growth curve and Ct value ≤40 as positive, corresponding to lower limits of detection from 143 to 875 copies/m^3^ in our air samples.

### Statistical methods

For each virus, we divided all the air sampling occasions into either of the two categories, whether (Scenario 1) there was at least one patient in the room who was tested positive for the infection, or (Scenario 2) none of the patients in the room was tested positive for the infection. We further divided the first category into two sub-categories, (Scenario 1a) the air sampler was placed where at least one of the patients in the two adjacent beds (“neighboring” patients) tested positive for the infection, regardless of whether any of the other patients in the room also tested positive or not, or (Scenario 1b) the air sampler was placed where both neighboring patients in the adjacent beds tested negative for the virus, but in the same room there were other patient(s) who tested positive. To identify any difference between the virus for each scenarios (1a, 1b, 2), we compared the frequency of occasions with virus RNA/DNA recovery in the air for each air size fractions (<1µm, 1-4µm and >4µm) by Chi-square test or Fisher’s exact test. We also translated Ct values in the original samples to copies/m^3^ air based on flow rates. All analyses were conducted with R version 4.0.5 (R Foundation for Statistical Computing, Vienna, Austria).

## RESULTS

We collected air samples on 44 occasions from December 2017 to January 2020. Since we used two samplers on each occasion, and there were three size fractions in each sampler, we obtained in total 264 air samples for laboratory analysis. Patients triggering the air sampling included 15 (34%) patients with RSV, 14 (32%) with AdV, 12 (27%) with PIV and 3 (7%) with influenza B virus. These target patients had been admitted for a mean of 3.5 days (interquartile range, IQR 2.0-4.0) prior to sampling, and this was similar across viruses: 2.9 days (IQR 2.0-3.3) for RSV, 3.2 (IQR 2.3-3.0) for AdV, 4.4 days (IQR 1.3-4.8) for PIV and 3.1 days (IQR 2.4-3.9) for influenza B. For the 44 air sampling occasions, the average measured room temperature, relative humidity, and CO_2_ (ppm) were 25°C (IQR 23-26), 63% (IQR 49-78) and 1003 ppm (IQR 786-1224), respectively (Table 1). On average, at least 3 visitors and a maximum of 9 healthcare workers were present in the room.

**Table 1.** Characteristic of sampling occasions with at least one patient positive for the corresponding respiratory virus infection (Scenario 1).

| Occasions which at least one patient in the room positive for the corresponding respiratory virus infection | **RSV**<br>**(n=24)** | **AdV**<br>**(n=18)** | **PIV**<br>**(n=16)** | **Influenza B**<br>**(n=4)** | **Influenza A**<br>**(n=8)** | **All occasions**<br>**(n=44)** |
| --- | --- | --- | --- | --- | --- | --- |
| Patients No. | 133 | 95 | 84 | 23 | 42 | 239 |
| CO<sub>2</sub> (ppm) (IQR) | 959 (772 - 1132) | 1048 (817 - 1242) | 1059 (883 - 1222) | 754 (668 - 840) | 922 (716 - 1084) | 1003 (786 - 1224) |
| Room temperature (°C) (IQR) | 24 (23 - 25) | 25 (23 - 26) | 25 (24 - 26) | 23 (23 - 23) | 25 (24 - 26) | 25 (23 - 26) |
| Relative humidity (%) (IQR) | 68 (50 - 86) | 63 (51 - 68) | 66 (55 - 71) | 42 (33 - 47) | 57 (47 - 59) | 63 (49 - 78) |
| Healthcare workers (HCWs) No. (range) | ≤9 (0 - 9) | ≤9 (0 - 9) | ≤3 (0 - 3) | ≤7 (0 - 4) | ≤9 (0 - 9) | ≤9 |
| Visitors No. (range) | ≥3 (3 - 10) | ≥3 (3 - 10) | ≥4 (4 - 10) | ≥4 (4 - 8) | ≥4 (4 - 10) | ≥3 |
| Patients conducted nebulizer therapy in the room No. (%) | 82/113 (73) | 66/95 (69) | 55/84 (65) | 10/23 (43) | 26/42 (62) | 145/239 (61) |
| ***Patients in the room who were tested positive for a particular respiratory virus No. (%)*** |  |  |  |  |  |  |
| RSV | 34 | 9 | 13 | 0 | 2 | 34 |
| AdV | 8 | 18 | 3 | 1 | 5 | 18 |
| PIV | 8 | 3 | 19 | 0 | 3 | 19 |
| Influenza B | 0 | 1 | 0 | 5 | 1 | 5 |
| Influenza A | 2 | 5 | 3 | 1 | 8 | 8 |

In total, there were 239 pediatric patients present in the rooms during the 44 sampling occasions. These 239 patients had an average age of 3.3 years old (IQR 0.9-4.5), with a mean hospital stay of 8 days (IQR 4-10) between admission and discharge. Nebulization therapy was frequently conducted, with 80% of our sampling occasions having at least 3 patients conducting nebulization therapy before or during the air sampling on the same day. Information on laboratory testing results was available for 222/239 (93%), including 110/239 (46%) who were positive for RSV, AdV, PIV, influenza B or influenza A (Table 1). From the 44 air sampling occasions we identified occasions which had at least one patient in the room who tested positive for a particular respiratory virus, including 24 (55%) for RSV, 18 (41%) for AdV, 16 (36%) for PIV, 4 (9%) for influenza B and 8 (20%) occasions for influenza A (Table 1).

We classified the 44 air sampling occasions into one of the three scenarios of whether there were patients with confirmed infection in the room for each respiratory virus, and present the frequency of occasions with respiratory virus recovery in the air at any of the three air size fractions (Table 2), or stratified by the three size fractions (Table 3), and viral load identified in each of the three air size fractions (Figure 1). The three scenarios were: *Scenario 1a* – occasions with neighboring patients positive for the corresponding RV infection; *Scenario 1b* – occasions with neighboring patients negative but other patients in the room positive for the corresponding RV infection; and *Scenario 2* – occasions where no patients in the room was positive for the corresponding respiratory virus infection. We also present the data in a subset of sampling occasions which the neighboring patients were the only patients in the room positive for the infection (Table 4). Separately, for Scenarios 1a and 1b, we also compared the respiratory virus recovery between whether the target patient had undergone nebulization or not during the air sampling (Table 5).

**Table 2.** Frequency of occasions with respiratory virus RNA/DNA detection in the air in any air size fractions for five respiratory viruses.

|  | RSV | AdV | PIV | Flu B | Flu A | p value |
| --- | --- | --- | --- | --- | --- | --- |
|  | No. of occasions positive / Total no. of occasions (%) |  |  |  |  |  |
| *Air sampling occasions* |  |  |  |  |  |  |
| Occasions which at least one patient in the room positive for the corresponding respiratory virus infection (Scenario 1a + 1b) | 24/44 (55) | 18/44 (41) | 16/44 (36) | 4/44 (9) | 8/44 (20) | - |
| *Respiratory virus RNA/ DNA recovery in the air* |  |  |  |  |  |  |
| Occasions with the corresponding respiratory virus RNA/DNA recovered in the air | 12/44 (27) | 10/44 (23) | 2/44 (5) | 12/44 (27) | 31/44 (70) | - |
| Respiratory virus recovery rate in occasions with neighboring patients positive for the corresponding virus infection (Scenario 1a) | 2/16 (13) | 6/16 (38) | 1/13 (8) | 2/4 (50) | 1/2 (50) | 0.1 |
| Respiratory virus recovery rate in occasions with neighboring patients negative but other patients in the room positive for the corresponding virus infection (Scenario 1b) | 4/8 (50) | 1/2 (50) | 0/3 (0) | 0/0 (0) | 4/6 (67) | 0.3 |
| Respiratory virus recovery rate in occasions where no patients in the room were positive for the corresponding virus infection (Scenario 2) | 6/20 (30) | 3/26 (12) | 1/28 (4) | 10/40 (25) | 26/36 (72) | <0.001 |

RSV: respiratory syncytial virus; AdV: adenovirus; PIV: parainfluenza virus.

**Table 3.**
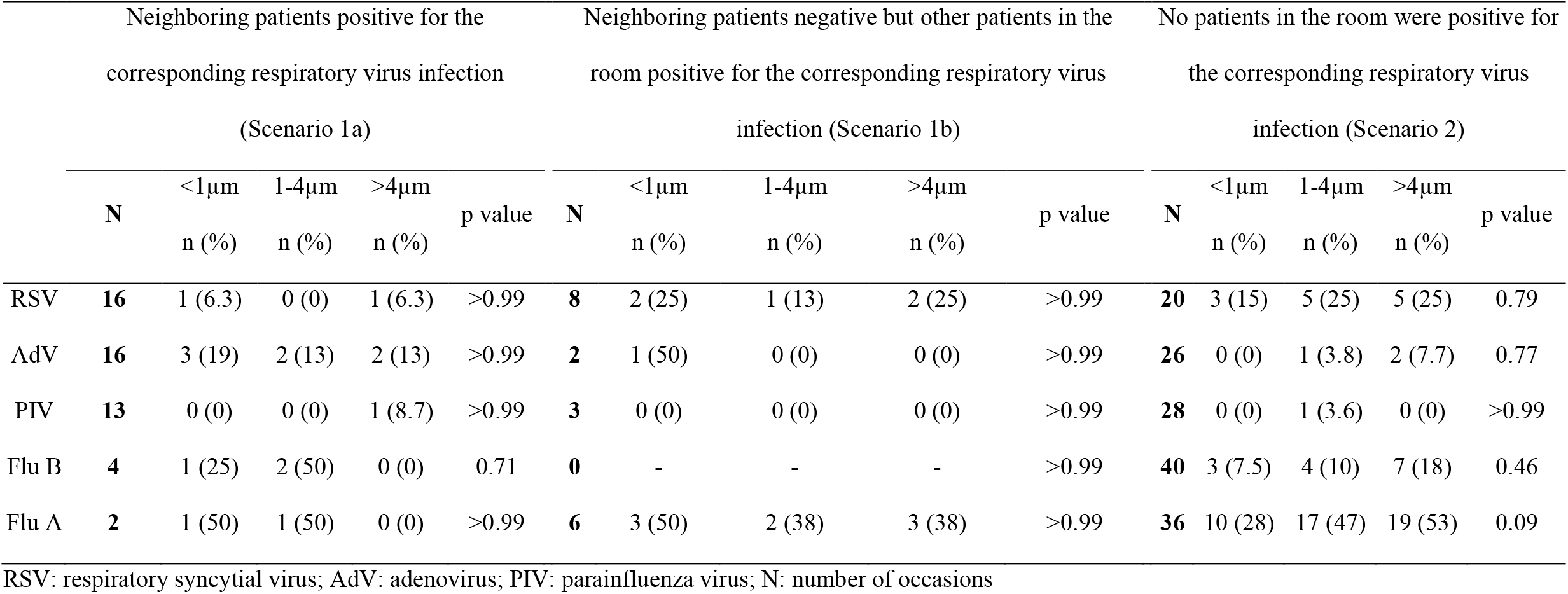
Frequency of occasions with respiratory virus RNA/DNA detection in the air in three air size fractions (<1µm, 1-4µm and >4µm) for five respiratory viruses.

|  | Neighboring patients positive for the<br>corresponding respiratory virus infection<br>(Scenario 1a) |  |  |  |  | Neighboring patients negative but other patients in the<br>room positive for the corresponding respiratory virus<br>infection (Scenario 1b) |  |  |  |  | No patients in the room were positive for<br>the corresponding respiratory virus<br>infection (Scenario 2) |  |  |  |  |
| --- | --- | --- | --- | --- | --- | --- | --- | --- | --- | --- | --- | --- | --- | --- | --- |
|  | N | <1µm<br>n (%) | 1-4µm<br>n (%) | >4µm<br>n (%) | p value | N | <1µm<br>n (%) | 1-4µm<br>n (%) | >4µm<br>n (%) | p value | N | <1µm<br>n (%) | 1-4µm<br>n (%) | >4µm<br>n (%) | p value |
| RSV | **16** | 1 (6.3) | 0 (0) | 1 (6.3) | >0.99 | **8** | 2 (25) | 1 (13) | 2 (25) | >0.99 | **20** | 3 (15) | 5 (25) | 5 (25) | 0.79 |
| AdV | **16** | 3 (19) | 2 (13) | 2 (13) | >0.99 | **2** | 1 (50) | 0 (0) | 0 (0) | >0.99 | **26** | 0 (0) | 1 (3.8) | 2 (7.7) | 0.77 |
| PIV | **13** | 0 (0) | 0 (0) | 1 (8.7) | >0.99 | **3** | 0 (0) | 0 (0) | 0 (0) | >0.99 | **28** | 0 (0) | 1 (3.6) | 0 (0) | >0.99 |
| Flu B | **4** | 1 (25) | 2 (50) | 0 (0) | 0.71 | **0** | - | - | - | >0.99 | **40** | 3 (7.5) | 4 (10) | 7 (18) | 0.46 |
| Flu A | **2** | 1 (50) | 1 (50) | 0 (0) | >0.99 | **6** | 3 (50) | 2 (38) | 3 (38) | >0.99 | **36** | 10 (28) | 17 (47) | 19 (53) | 0.09 |

RSV: respiratory syncytial virus; AdV: adenovirus; PIV: parainfluenza virus; N: number of occasions

**Table 4.** Frequency of occasions with respiratory virus RNA/ DNA detection in the air in three air size fractions (<1µm, 1-4µm and >4µm) for five respiratory viruses, restricted to occasions which the neighboring patients were the only patients in the room positive for the infection.

|  | N | Closer NIOSH sampler |  |  |  | Further NIOSH sampler |  |  |  |
| --- | --- | --- | --- | --- | --- | --- | --- | --- | --- |
|  |  | Any fraction | <1µm | 1-4µm | >4µm | Any fraction | <1µm | 1-4µm | >4µm |
|  |  | n (%) | n (%) | n (%) | n (%) | n (%) | n (%) | n (%) | n (%) |
| RSV | **12** | 1 (8) | 1 (8) | 0 (0) | 0 (0) | 1 (8) | 1 (8) | 0 (0) | 0 (0) |
| AdV | **16** | 4 (25) | 1 (6) | 1 (6) | 2 (13) | 5 (31) | 2 (13) | 1 (6) | 2 (13) |
| PIV | **12** | 0 (0) | 0 (0) | 0 (0) | 0 (0) | 0 (0) | 0 (0) | 0 (0) | 0 (0) |
| Influenza B | **3** | 1 (33) | 1 (33) | 1 (33) | 0 (0) | 1 (33) | 1 (33) | 0 (0) | 0 (0) |
| Influenza A | **2** | 1 (50) | 1 (50) | 1 (50) | 0 (0) | 1 (50) | 0 (0) | 1 (50) | 0 (0) |

RSV: respiratory syncytial virus; AdV: adenovirus; PIV: parainfluenza virus; N: number of occasions

**Table 5.**
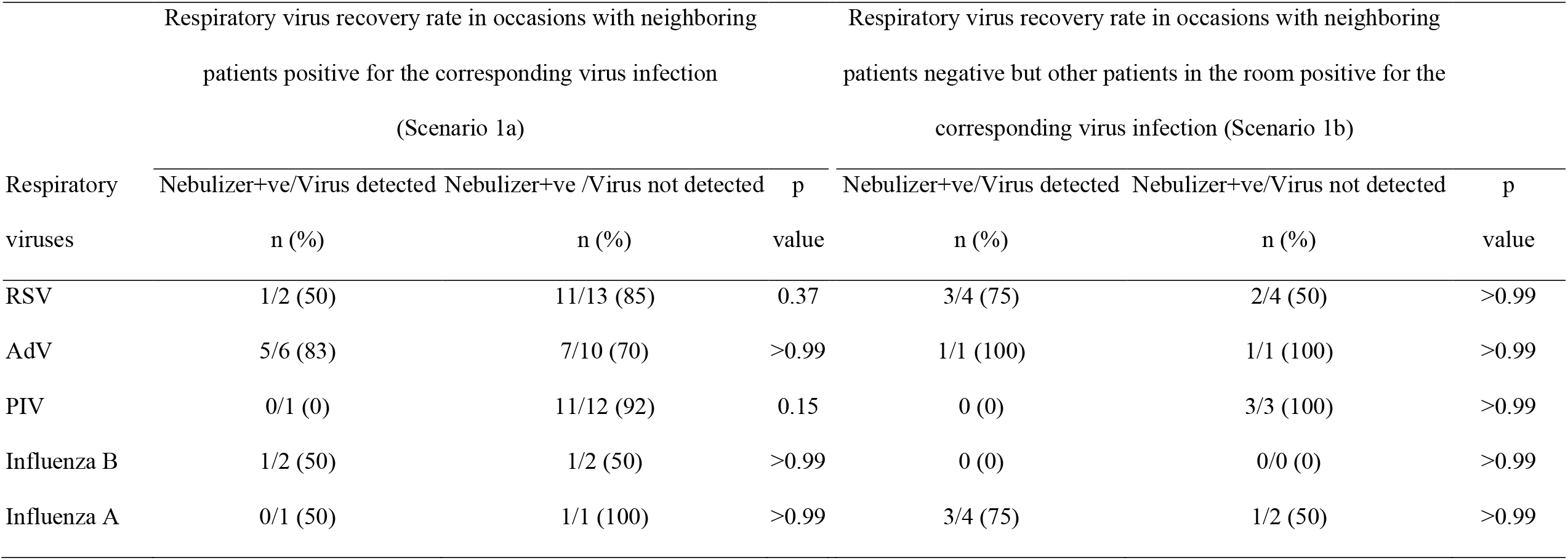
Frequency of respiratory virus detections in the subset of sample collections when nebulizer therapy was being performed for patients with confirmed respiratory virus infection of the corresponding type.

**Figure 1.**
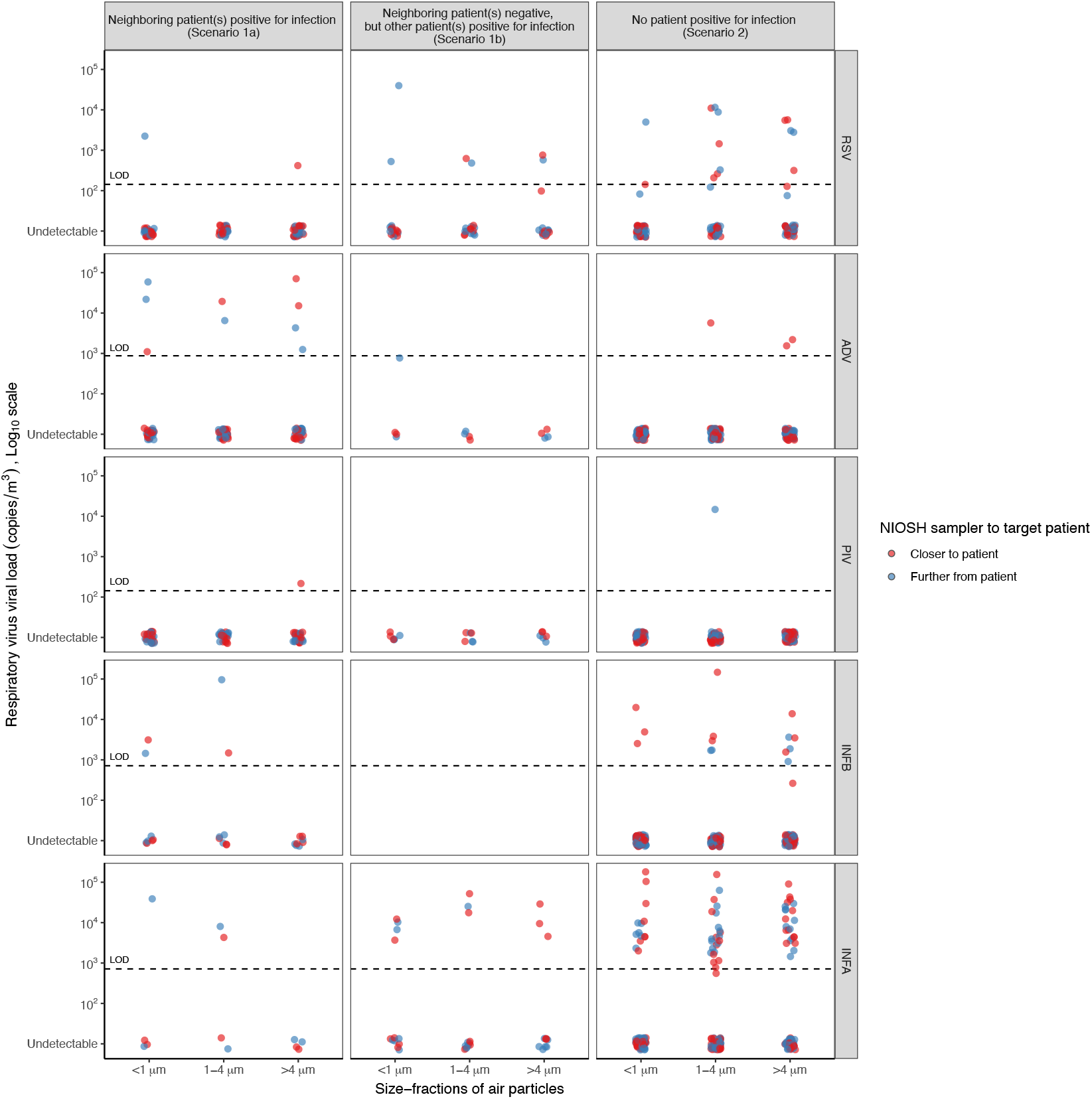
Viral load of respiratory virus RNA/ DNA recovered in the air in pediatric patient rooms in three air size fractions (<1µm, 1-4µm and >4µm) for five respiratory viruses. Respiratory viruses tested including respiratory syncytial virus (RSV), adenovirus (AdV), parainfluenza virus (PIV), influenza B virus and influenza A virus. For each respiratory virus (rows), the air sampling occasions are categorized into three scenarios (columns): *Scenario 1a* – occasions with neighboring patients positive for the corresponding virus infection; *Scenario 1b* – occasions with neighboring patients negative but other patients in the room positive for the corresponding virus infection; and *Scenario 2* – occasions where no patients in the room was positive for the corresponding virus infection. In each occasion we placed two air samplers in the room, including one sampler placed next to (red) and another sampler (blue) placed further away from the target patent. Viral load is expressed as viral copies per m^3^ air sampled, and the limit of detection (LOD) are 143 copies/m^3^ for RSV and PIV, 714 copies/m^3^ for influenza and 875 copies/m^3^ for AdV, respectively.

Among all 44 air sampling occasions, influenza A gene copies was more frequently detected in the air (70%), followed by influenza B (27%), RSV (27%) and AdV (23%), and PIV (5%) was rarely detected (Table 2). However, all respiratory viruses were recovered in the air in patient rooms even in the absence of known confirmed patients in the rooms, with significant difference in the detection between respiratory viruses, and observed most frequent recovery for influenza A (72%), RSV (30%) and influenza B (25%) and less for AdV (12%) and PIV (4%) (Scenario 2). If restricting to air sampling occasions where at least one patient in the room who had confirmed infection for the corresponding virus, we observed virus recovery in the air for all RVs when there were infected patients in close proximity (Scenario 1a); and even virus recovery when there were no known infected patients in close proximity for influenza A (4/6 occasions, 67%), RSV (4/8 occasions, 50%) and AdV (1/2 occasion, 50%), while the study was underpowered for the other two viruses (Scenario 1b).

Viral gene copies were recovered in all air size fractions for all respiratory viruses except PIV (Table 3, Figure 1). Despite the presence of confirmed patients in close proximity (Scenario 1a), we observed only small proportions (mostly <20%) of occasions with virus recovery for RSV, AdV and PIV, and no observed difference between size fractions. Virus recovery in all size fractions was observed when known infected patients were not present in close proximity but in the same room for RSV and INFA (Scenario 1b). Even when there were no known patients with the confirmed infection in the room (Scenario 2), we could recover viral gene copies in all size fractions for influenza A virus, RSV and INFB, furthermore for influenza A apparently there were higher proportions of virus recovery in the larger particles (Table 3). When restricting to air sampling occasions which the neighboring patients were the only patients in the room positive for the infection, viral gene copies were recovered in less than 15% of all fractions for RSV and AdV, and none of all fractions for PIV (Table 4).

More than 60% of the 145/239 pediatrics patients conducted nebulizer therapy during the 44 air sampling occasions, and nebulizers were used by at least 3 patients during air sampling for 34/44 (80%) of occasions. We did not find an association between the frequency of nebulizer use and the detection of respiratory viruses in our air samples (Table 5).

## DISCUSSION

We conducted a 2-year air sampling study in pediatric rooms before the COVID-19 pandemic and tested for five of the most common respiratory viruses in children. We had expected the detection of respiratory virus RNA/DNA to occur more frequently when we placed the sampler next to a patient with that viral infection and less likely on other occasions, but this was not always the case (Table 4). We frequently detected influenza A virus in our samples even when there was no patient in the room with confirmed influenza A virus infection, consistent with the findings in our earlier study in the same setting which focused on influenza [22], which was expected because influenza A was circulating in the community during the air sampling, and Xie et al. reported that increased crowdedness was associated with a higher frequency of detection of virus-laden particles [5]. In the present study, we showed that this also occurred to a lesser extent with influenza B virus and RSV (Table 2).

These findings are compatible with other observations of influenza virus [26,27] or RSV [11,19,28] detection in the air far from the presumed source in healthcare settings. Aintablian et al. frequently identified RSV RNA in pediatric patient rooms at distances as far as 7m from the patient bedside [19]. In our study, 30% of RSV was recovered in the air in patient rooms even in the absence of known confirmed patients in the rooms, while Grayson et al. reported detection of RSV in air samples on 4/8 sampling days without RSV confirmed patients in the pediatric exam rooms and most of the RSV-laden particles were larger than 4µm [10]. The present study suggests that respiratory viruses recovered from the air in patient rooms may be from patients in close proximity, from other more distant patients in the same room, or even perhaps from other sources in the hospital such as patients in other rooms, healthcare workers or visitors [11].

The less frequent detection of AdV and especially rarely for PIV in our air samples might perhaps be indicative of a lower chance of transmission via the aerosol route for these respiratory viruses. We observed that AdV was more frequently recovered in the air when the sampler was placed next to an infected patient but less when there were no infected patients in close proximity (Figure 1). However, AdV was frequently detected in studies in pediatric emergency rooms in Taiwan [13] and Brazil [16], and general pediatric wards in Singapore [29]. There were also occasional reports of AdV outbreaks in healthcare settings such as a children’s hospital [30] and a pediatric care facility [31]. We detected AdV in all size fractions, mostly from patients with confirmed AdV infection at admission, suggesting an opportunity for infection control through rapid diagnosis at admission if nosocomial AdV outbreaks occur.

Rare detection of PIV in the air from the environment was also reported in most of other studies [17,32,33]. Mclean et al. used Anderson samplers and detected PIV in air samples collected from 1/30 children sampled [17]. Goyal et al. reported the presence of PIV in 1/64 used ventilation filters in public buildings [32]. Another study in Korea reported a total detection rate of 0.5% in air samples collected in a community setting [33]. In contrast to these evidence, Gralton et al. identified PIV RNA in exhaled breath or cough samples collected from 13/53 (26%) infected children and adults [18], and animal studies showed that airborne transmission of murine PIV (Sendai virus) was 86-100% efficient [34] and short-range airborne transmission was warranted [35]. More studies, perhaps on the stability of PIV in aerosols, may help to explain the discrepancy.

The potential for aerosol generating procedures to increase the risk of nosocomial transmission have been suggested [36]. In our study patients were often prescribed nebulizer therapy, but we did not find an association between nebulizer therapy and the detection rates of virus (Table 5). This may be because coughing of patients in the ward had already generated virus-laden aerosols which were detected in our samplers [18], whether or not aerosol generating procedures were also used.

The high levels of viral gene copies at the scale of 10^3^-10^5^ copies/m^3^ air detected in our study in 5-bed pediatric patient rooms, particularly for influenza A and B viruses and RSV (Figure 1), are similar to another study in pediatric outpatient clinics [13] but contrast to the low levels reported in the air in other studies in adult patient rooms [37]. Previously we reported a ventilation rate of 1.5 air changes per hour in a similar but unoccupied patient room in the same hospital [22], and low ventilation may allow the accumulation of respiratory viruses in the air to higher concentrations. Alternatively, patients for whom we placed the air samplers nearby may be earlier in their disease course, so that viral shedding may tend to be higher [38], and children may shed more virus than adults in general. However, our study did not show higher virus recovery when there were infected patients in close proximity (Table 4). We also have limited understanding on the viral shedding kinetics and their relationship with symptom onset for many respiratory viruses.

Our study has several limitations. First, we did not culture virus among the recovered air samples, since we expected low isolation rate based on our previous study on influenza in the same setting [22]. Second, we were not able to identify the source of generation of virus detected in the air nor transmission via the aerosol route among children. It is possible that infected staff or visitors who visited the room during the air sampling were responsible for some of the virus detected in the air. Virus sequencing could be used in future studies to help identify the source of virus detections. Third, we did not measure the ventilation rate during the sampling occasions, but referred to data from our previous study [22], but human and other environmental movement such as opening and closing windows, doors, and drying racks might affect ventilation rates.

In conclusion, influenza A/B viruses and RSV were frequently detected in the air in pediatric rooms, including frequently in rooms where no patients in the room tested positive for the corresponding virus infection. PIV was rarely detected. We did not find an association of virus detection with the use of nebulizer therapy. Our results indicate the potential benefits of improving personal protective equipment for staff and visitors, and/or improving indoor ventilation, both within patient rooms with known infected patients or in all healthcare settings in general. The latter may be achieved by increasing air changes through natural or mechanical ventilation, or perhaps air disinfection with ultraviolet light [4].

## Data Availability

All data produced in the present study are available upon reasonable request to the authors

## ACKNOWLEDGEMENTS

We wish to acknowledge Weijia Xiong for statistical support, colleagues from the Department of Infection Control and the State Key Laboratory of Respiratory Diseases at the First Affiliated Hospital of Guangzhou Medical University for technical support; and Dr William Lindsley from the US Centers for Disease Control and Prevention for providing the air samplers. This work was supported in part by the Theme-based Research Scheme from the Research Grants Council of the Hong Kong Special Administrative Region, China [Project No. T11-712/19-N], the Guangzhou Medical University High-level University Clinical Research and Cultivation Program [Guangzhou Medical University released [2017] No. 160]; the Guangzhou Medical University High-level University Innovation Team Training Program [Guangzhou Medical University released [2017] No. 159]; and the Science Research Project of the Guangdong Province, China [Grant No. 2016A050503047].

